# Seroprevalence of SARS-CoV-2 Infection in Cincinnati Ohio USA from August to December 2020

**DOI:** 10.1101/2021.03.11.21253263

**Authors:** Greg Davis, Allen J. York, Willis Clark Bacon, Lin Suh-Chin, Monica Malone McNeal, Alexander E. Yarawsky, Joseph J. Maciag, Jeanette L. C. Miller, Kathryn C. S. Locker, Michelle Bailey, Rebecca Stone, Michael Hall, Judith Gonzalez, Alyssa Sproles, E. Steve Woodle, Kristen Safier, Kristine A. Justus, Paul Spearman, Russell E. Ware, Jose A. Cancelas, Michael B. Jordan, Andrew B. Herr, David A. Hildeman, Jeffery D. Molkentin

## Abstract

The world is currently in a pandemic of COVID-19 (Coronavirus disease-2019) caused by a novel positive-sense, single-stranded RNA β-coronavirus referred to as SARS-CoV-2. Fortunately, most infected individuals recover and are then resistant to re-infection for a period, indicating that a vaccination approach can be successful. Elucidation of rates of past SARS-CoV-2 infection within select regions across the United States of America (USA) will help direct vaccination efforts and together will inform our approach towards achieving herd immunity. Here we investigated rates of SARS-CoV-2 infection in the greater Cincinnati, Ohio, USA metropolitan area from August to December 2020, just prior to initiation of the national vaccination program. Examination of 9,550 adult blood donor volunteers for serum IgG antibody positivity against the SARS-CoV-2 Spike protein showed an overall prevalence of 8.40%, measured as 7.56% in the first 58 days of this time frame, versus a significant increase to 9.24% in the last 58 days, and a final rate of 12.86% in December 2020. Approximately 56% of Spike seropositive individuals also had immunoreactivity against the receptor binding domain (RBD) within the Spike protein, which is associated with viral neutralization. Males and females in the Cincinnati area showed nearly identical rates of past infection, and rates among Hispanics, African Americans and Caucasians were not significantly different. Interestingly, donors under 30 years of age had the highest rates of past infection, while those over 60 had the lowest. Geographic analysis showed that the West side of Cincinnati had a rate of 9.63% versus 8.13% on the East side (demarcated by Interstate-75), while the adjoining area of Kentucky was 7.04% (as demarcated by the Ohio River). These results among healthy blood donors will be critical in calculating the time needed to achieve regional herd immunity in conjunction with the national vaccination campaign.

## Introduction

Severe acute respiratory syndrome coronavirus 2 (SARS-CoV-2), the agent responsible for Coronavirus disease 2019 (COVID19) was first identified in Wuhan, China in late 2019 (1,2), which thereafter spread across the globe resulting in the current pandemic. In the United States of America (USA) more than 525,000 deaths have been attributed to this virus as of March 2021, with approximately 29 million total documented cases of infection, of which over 980,000 were in the State of Ohio (3). Current efforts to curtail and possibly end the pandemic are largely dependent on the vaccine program underway across the USA and the world, with the goal of achieving herd immunity and dramatic reductions in viral spreading (4,5). Another important consideration in achieving herd immunity and the eventual end to the current pandemic is quantifying background rates of previous infection within the population (4). In general, previously infected individuals are resistant to new infection for a period, and/or they appear to have a reduced severity of disease if re-infected (6). Thus, characterizing levels of previous infection within communities across the USA will inform progression toward herd immunity with optimized vaccination strategies.

SARS-CoV-2 contains a large surface facing glycoprotein called Spike (S, ∼190 kDa) that facilitates binding of the virus to the receptor angiotensin converting enzyme 2 (ACE2) on host cells, which after proteolytic cleavage of S mediates viral cellular involution and infection (7,8). The receptor-binding domain (RBD, 30 kDa) is part of the S protein that directly interacts with ACE2, and antibodies against the RBD region can mediate neutralization and protection from viral infection (7,8). The S protein is also a primary component of immunogenicity for the host response against the virus that can produce some degree of lasting immunity (7). Thus, it is not surprising that a primary strategy for vaccine development against SARS-CoV-2 involves ectopic expression of the S protein to generate host specific neutralizing antibodies, particularly those directed against the RBD region (7,8).

Rates of seropositivity for SARS-CoV-2 in voluntary blood donors in the Greater Cincinnati Metropolitan Area (GCMA) of Ohio USA were examined from August 13th - December 8th of 2020, just prior to the beginning of the nationwide vaccination program. A modified serological enzyme-linked immunosorbent assays (ELISA) assay developed by Krammer and colleagues (9,10) was implemented to examine 9,550 individuals of age rage 16-91 years old for SARS-CoV-2 S protein IgG antibodies to quantify rates of prior infection in the GCMA. All positives were assayed separately for concomitant IgG sero-reactivity against the RBD portion of the S protein. A cross-section of the positive ELISA data was further examined using the Luminex immunodetection platform for potential cross-reactivity with the 4 endemic human cold-causing coronaviruses (HCoV- 229E, -NL63, -OC43, and -HKU1) (11,12). Since some individuals donated blood more than once over this 5-month period, rates of decline in S protein antibody levels were documented, suggesting some potential reduction in immunity. Finally, rates of S protein seropositivity in the GCMA due to ethnicity, gender, and geographic subregions were examined.

## Results

To document rates of previous SARS-CoV-2 infection in the GCMA leading up to the onset of the national vaccination program, blood samples from randomly presenting volunteer donors through the Hoxworth Blood Center were collected and analyzed for antibodies against the S protein by ELISA, and positives were also analyzed for RBD antibodies in a separate ELISA. Exactly 10514 samples were collected and processed from August 13th through December 8th of 2020, which represented 9550 unique donors and we determined that 802 were positive for S protein antibodies, for an overall prevalence of 8.40% (Table 1). The accuracy of this reported rate of past infection is dependent on how the laboratory ELISA was implemented and verified, which is detailed in the Methods section. The ELISA itself was based on an established protocol and set of biological reagents described by Krammer and colleagues (9,10), which was granted Emergency Use Authorization by the US Food and Drug Administration. However, a more robust protein production system based in expi-CHO cells was used to generate S and RBD proteins, which yielded greater amounts than observed using expi293 cells described by Krammer and colleagues (9,10) (Supplemental Figure 1). An improved purification protocol with quality control analytics was also instituted (Supplemental Figure 1) to ensure the purity of the preparations and that the S protein was in the trimeric configuration (Supplemental Figure 1). However, the ELISA-based reactivity of S protein antigen generated in expiCHO cells or expi293 cells was essentially identical (data not shown). The ELISA positivity cut-off was set as 3-standard deviations above background, calculated with serum samples from 2019 before the onset of the pandemic. The actual experimental value from the ELISA was 0.4039 for S protein IgG antibody and 0.4826 for RBD as optical density (OD) units, so that readings higher than these values were interpreted as positive (Table 2 and Figure 1). Importantly, this was a blinded study in which SARS-CoV-2 testing was performed post-hoc on de-identified specimens and did not modify or influence donor recruitment, eligibility or deferral for this donation or subsequent donations.

**Table 1.**
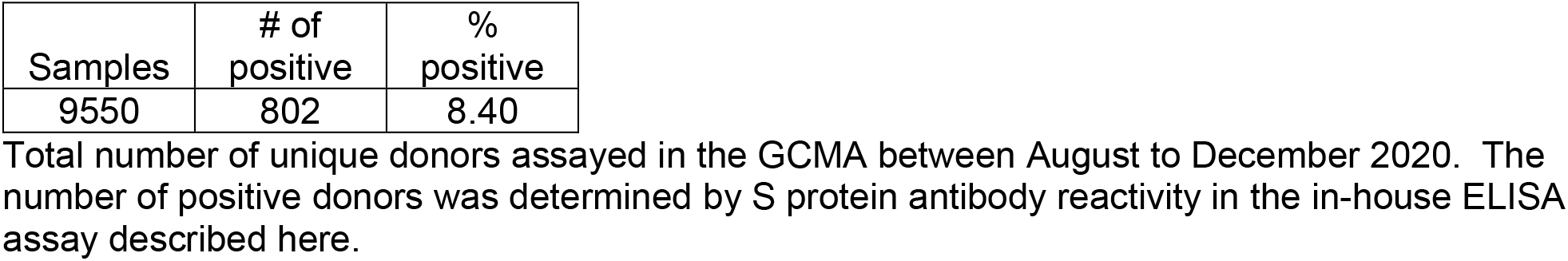
Donor S protein IgG antibody positivity rate in Hoxworth blood samples collected from August 13, 2020 to December 8, 2020.

| Samples | # of positive | % positive |
| --- | --- | --- |
| 9550 | 802 | 8.40 |

Total number of unique donors assayed in the GCMA between August to December 2020. The number of positive donors was determined by S protein antibody reactivity in the in-house ELISA assay described here.

**Table 2.** Mean S and RBD raw ELISA OD values from human serum prior to 2020.

|  | Mean of negative controls | Standard deviation | Mean + 3 standard deviations |
| --- | --- | --- | --- |
| Spike OD | 0.1284 | 0.09185 | 0.4039 |
| RBD OD | 0.1482 | 0.1115 | 0.4826 |

The final ELISA raw data for antibody reactivity against S or RBD proteins, as measured in OD values at 492 nm in an automated spectrophotometric plate reader. The assay standard deviation was determined with 60 and 53 negative control serum samples taken before the COVID19 pandemic in 2019 for S and RBD, respectively. Three standard deviation units was added to the negative control value in setting the assay threshold positivity value shown for S and RBD.

**Figure 1.**
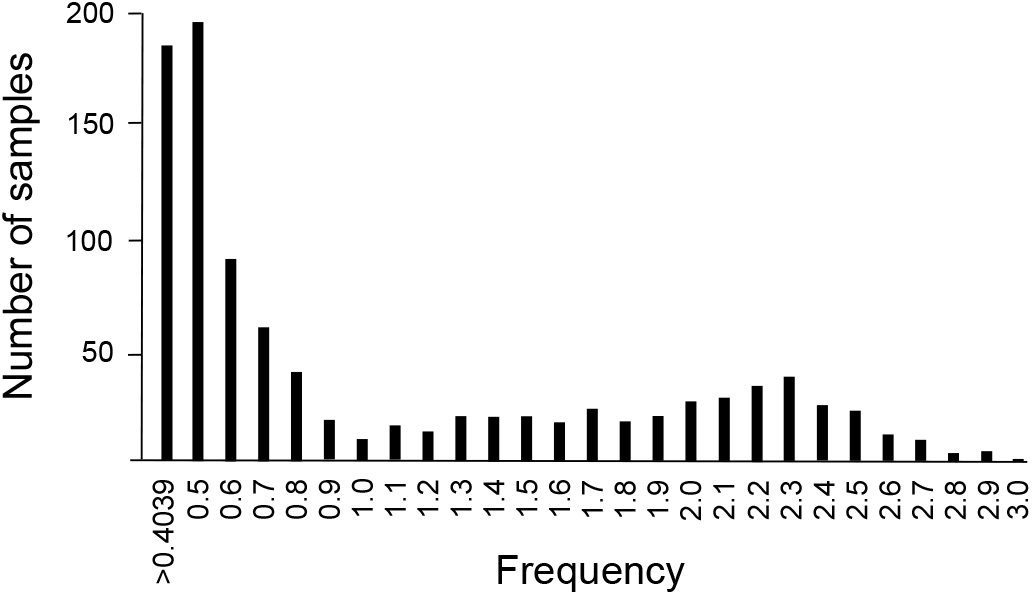
Histogram frequency of positive S protein ELISA OD values from 0.4039 to 3.000. The 802 Spike positive samples were grouped in bins of 0.1 OD units with the initial bin starting at 0.4039, the minimum positive Spike OD value. The maximum Spike OD of all samples was 2.998.

All 802 of 9550 donors with antibodies against the S protein were also evaluated for antibodies against the RBD in separate ELISAs, which showed that 446 were positive for both, a rate of 55.61% (Table 3). However, because the RBD region is ∼85% smaller than the entire S protein, there are fewer total antibodies with specificity to RBD compared with S protein, explaining the reduced sensitivity of the RBD ELISA and likely why only ∼56% concordance was observed (Table 3). Indeed, a sub-analysis of the 802 positive donors for S protein based on ELISA intensity was performed, and the highest 1/3rd OD values (2.0-3.0) showed 97.3% positivity for RBD, while the lowest 1/3rd of OD values (0.4039-1.0) only showed 21.3% positivity for RBD (Table 3). Thus, those donors with the lowest levels of total S protein antibodies were less likely to have detectable RBD antibodies simply based on sensitivity issues.

**Table 3.** S and RBD positivity rates analyzed over 3 ranges of S protein antibody OD values.

|  | # of Spike Pos | # of RBD Pos | % RBD and Spike positive |
| --- | --- | --- | --- |
| All positives: | 802 | 446 | 55.61 |
| Spike OD range |  |  |  |
| 0.4039 to 1.000 | 409 | 87 | 21.27 |
| 1.001 to 2.000 | 205 | 176 | 85.85 |
| 2.000 to 3.000 | 188 | 183 | 97.34 |

Exactly 802 donors that were positive for S protein antibodies were also assessed for RBD ELISA based antibody positivity, and while only 55.61% were positive for both S and RBD, this rate depended on the strength of the S protein antibody levels over the 3 different OD ranges shown. Thus, those donors with high S protein antibodies were much more like to be positive for RBD antibodies.

The Luminex immunodetection platform was also employed to examine potential cross-reactivity with the endemic 4 human cold-causing coronaviruses that might influence S protein or RBD reactivity in the ELISAs, as well as to confirm the validity and sensitivity of the S protein ELISA (Table 4). We selected a group of 11 donor samples that were highly positive for S protein and RBD by ELISA for comparative analysis in the Luminex platform, which as expected showed a rate of 100% positivity for S, RBD and Nucleocapsid (N) protein from SARS-CoV2, but not S protein from MERS-CoV or SARS-CoV-1, which further validates the ELISA protocol (Table 4). Eleven highly characterized negative controls from the S protein ELISA were also examined and these same samples were 100% negative for S protein, RBD and N protein in the Luminex assay (Table 4). To examine the sensitivity of the S protein ELISA we also analyzed 57 donor samples that were RBD negative and relatively low in overall S protein reactivity, and in this group 67% confirmed in the Luminex assay for S protein, and 23% were positive for RBD, but only one sample was positive for N protein (Table 4). These results with the Luminex assay support the validity of the ELISA but also suggest that the ELISA is more sensitive than the Luminex platform in detecting S protein.

**Table 4.** Comparison of S protein ELISA versus Luminex immunodetection of S protein, RBD, N and status of the 4 hCoVs and MERS and SARS-CoV-1.

|  | **ELISA** |  | **Luminex** |  |  |  |  |  |  |  |  |
| --- | --- | --- | --- | --- | --- | --- | --- | --- | --- | --- | --- |
| Negative controls (sample numbers) | Spike protein OD | RBD protein OD | SARS-CoV-2 Spike | SARS-CoV-2 Spike RBD | SARS-CoV-2 Nucleocapsid Protein | HCoV-229E Spike S1 | HCoV-HKU1 Spike S1 | HCoV-NL63 Spike S1 | HCoV-OC43 Spike S1 | MERS-CoV Spike S1 | SARS-CoV Spike S1 |
| N1 | 0.132 | 0.312 | 16.5 | 25.5 | 28.5 | 752.3 | 427.5 | 681.5 | 1451.5 | 18.5 | 25.5 |
| N2 | 0.159 | 0.196 | 39 | 4.5 | 2 | 1480.8 | 78.5 | 352.5 | 287.5 | 1.5 | 17.5 |
| N3 | 0.138 | 0.136 | 19 | 10.5 | 11 | 3016.3 | 539.5 | 538 | 1343.5 | 5.5 | 16.5 |
| N4 | 0.127 | 0.11 | 49 | 26.5 | 1091.5 | 844.3 | 1322.5 | 90 | 170.5 | 7.5 | 22.5 |
| N5 | 0.056 | 0.018 | 30.5 | 3.5 | 30.5 | 726.3 | 525.5 | 462.5 | 543.5 | 1.5 | 6.5 |
| N6 | 0.251 | 0.21 | 29 | 57 | 73.5 | 3204.8 | 257.5 | 433.5 | 581 | 43.5 | 45 |
| N7 | 0.178 | 0.178 | 17 | 41 | 45.5 | 1494.8 | 253.5 | 777.5 | 213 | 31.5 | 33.5 |
| N8 | 0.203 | 0.181 | 113 | 38 | 17.5 | 613.3 | 835.5 | 287.5 | 545.5 | 10 | 13.5 |
| N9 | 0.128 | 0.12 | 74.5 | 12.5 | 15 | 1419.8 | 696 | 412 | 293.5 | 3.5 | 12.5 |
| N10 | 0.152 | 0.207 | 61 | 25.5 | 19.5 | 2794.8 | 456.5 | 938.5 | 506 | 7.5 | 14.5 |
| N11 | 0.089 | 0.185 | 14 | 14.5 | 8.5 | 1496.8 | 548.5 | 463.5 | 596.5 | 3.5 | 10.5 |
|  | **ELISA** |  | **Luminex** |  |  |  |  |  |  |  |  |
| Positive Spike and Positive RBD Samples | Spike protein OD | RBD protein OD | SARS-CoV-2 Spike | SARS-CoV-2 Spike RBD | SARS-CoV-2 Nucleo-capsid Protein | HCoV-229E Spike S1 | HCoV-HKU1 Spike S1 | HCoV-NL63 Spike S1 | HCoV-OC43 Spike S1 | MERS-CoV Spike S1 | SARS-CoV Spike S1 |
| B1 | 2.904 | 2.991 | 21290.3 | 8917.7 | 5821.3 | 260.5 | 320.3 | 768.3 | 81.7 | 21.7 | 128.3 |
| B3 | 2.111 | 2.701 | 14502.8 | 4787.2 | 4541.3 | 3033 | 1447.3 | 403.3 | 1511.2 | 7.7 | 138.3 |
| B4 | 2.637 | 3.020 | 24563.3 | 11541 | 7229.3 | 3173 | 1588.3 | 477.3 | 583.2 | 11.7 | 100.3 |
| B5 | 3.000 | 2.944 | 16272.3 | 6672.2 | 4689.3 | 2183 | 1399.3 | 593.8 | 360.2 | 7.7 | 44.8 |
| B6 | 2.218 | 3.125 | 25727.3 | 13068 | 6875.8 | 2881.5 | 287.3 | 1614.3 | 755.7 | 8.7 | 61.3 |
| B7 | 1.380 | 3.049 | 24344 | 8703 | 7509 | 3266 | 970 | 1208 | 1324.5 | 11 | 106 |
| B8 | 1.432 | 3.132 | 25231.3 | 10711 | 6537.3 | 856 | 821.3 | 626.3 | 1900.2 | 23.7 | 103.3 |
| B9 | 2.019 | 2.952 | 16536.8 | 6820.7 | 5045.3 | 4267.5 | 460.8 | 2000.3 | 1277.7 | 10.2 | 153.3 |
| B10 | 2.643 | 3.042 | 27812.8 | 17379 | 9306.3 | 1560 | 1060.3 | 1241.3 | 406.2 | 9.7 | 403.3 |
| B11 | 2.701 | 2.662 | 23273.8 | 9145.7 | 14334.3 | 3689 | 332.3 | 1295.8 | 598.7 | 21.7 | 259.3 |

| Sample<br>number <1.0<br>for Spike,<br>RBD neg | ELISA |  | Luminex |  |  |  |  |  |  |  |  |
| --- | --- | --- | --- | --- | --- | --- | --- | --- | --- | --- | --- |
|  | Spike<br>protein<br>OD | RBD<br>protein<br>OD | SARS-CoV-2<br>Spike | SARS-CoV-2<br>Spike RBD | SARS-CoV-2<br>Nucleocapsid<br>Protein | HCoV-229E<br>Spike S1 | HCoV-HKU1<br>Spike S1 | HCoV-NL63<br>Spike S1 | HCoV-OC43<br>Spike S1 | MERS-CoV<br>Spike S1 | SARS-CoV1<br>Spike S1 |
| C1 | 0.506 | 0.159 | 35 | 47 | 78 | 1431 | 3386 | 593 | 2136 | 4 | 99 |
| C2 | 0.508 | 0.097 | 352 | 13 | 40 | 5246 | 1581 | 342 | 2480 | 3 | 13 |
| C3 | 0.51 | 0.133 | 47 | 41 | 203 | 438 | 3263 | 183 | 3974 | 4 | 84 |
| C4 | 0.51 | 0.258 | 300 | 13 | 24 | 505 | 405 | 82 | 838 | 2 | 15 |
| C5 | 0.513 | 0.169 | 431 | 15 | 48 | 1801 | 1455 | 677 | 4787 | 2 | 20 |
| C6 | 0.516 | 0.134 | 243 | 11 | 12 | 2062 | 797 | 138 | 1793 | 2 | 13 |
| C7 | 0.553 | 0.104 | 430 | 32 | 470 | 947 | 443 | 356 | 1160 | 13 | 28 |
| C8 | 0.561 | 0.065 | 1297 | 16 | 21 | 776 | 1873 | 741 | 2410 | 4 | 20 |
| C9 | 0.563 | 0.146 | 578 | 20 | 34 | 5654 | 3184 | 1058 | 5784 | 6 | 22 |
| C10 | 0.566 | 0.234 | 450 | 13 | 30 | 3929 | 597 | 289 | 1405 | 8 | 11 |
| C11 | 0.571 | 0.043 | 343 | 18 | 32 | 4008 | 1637 | 698 | 4811 | 3 | 19 |
| C12 | 0.574 | 0.222 | 1086 | 11 | 24 | 1004 | 1753 | 816 | 2718 | 4 | 16 |
| C13 | 0.579 | 0.298 | 44 | 25 | 60 | 1126 | 796 | 568 | 1911 | 2 | 14 |
| C14 | 0.581 | 0.191 | 580 | 7 | 41 | 659 | 1517 | 351 | 2412 | 2 | 64 |
| C15 | 0.587 | 0.255 | 213 | 11 | 18 | 3960 | 3817 | 667 | 3054 | 2 | 3 |
| C16 | 0.597 | 0.313 | 59 | 20 | 42 | 1292 | 1934 | 319 | 1182 | 5 | 21 |
| C17 | 0.601 | 0.309 | 2517 | 423 | 363 | 267 | 1089 | 73 | 2073 | 4 | 88 |
| c20 | 0.602 | 0.284 | 36 | 83 | 95 | 1954 | 3003 | 258 | 4087 | 6 | 57 |
| C21 | 0.613 | 0.319 | 25 | 24 | 51 | 3276 | 2953 | 206 | 547 | 3 | 18 |
| C22 | 0.614 | 0.173 | 413 | 35 | 49 | 3405 | 2164 | 551 | 2615 | 4 | 44 |
| C23 | 0.615 | 0.225 | 17 | 6 | 22 | 3331 | 206 | 406 | 1701 | 3 | 12 |
| C24 | 0.621 | 0.173 | 2352 | 1552 | 5752 | 218 | 1205 | 66 | 3895 | 8 | 124 |
| C25 | 0.629 | 0.342 | 25 | 12 | 27 | 1690 | 2301 | 640 | 149 | 4 | 22 |
| C26 | 0.636 | 0.08 | 276 | 132 | 249 | 750 | 636 | 304 | 639 | 1 | 179 |
| C27 | 0.639 | 0.337 | 582 | 190 | 502 | 4874 | 3450 | 240 | 3941 | 5 | 389 |
| C28 | 0.641 | 0.055 | 29 | 7 | 22 | 824 | 313 | 296 | 157 | 3 | 11 |
| C29 | 0.642 | 0.15 | 223 | 24 | 68 | 2828 | 2071 | 376 | 1308 | 2 | 16 |
| C30 | 0.645 | 0.22 | 156 | 102 | 240 | 3734 | 1090 | 245 | 2153 | 11 | 129 |
| C31 | 0.671 | 0.299 | 7 | 11 | 27 | 932 | 352 | 332 | 351 | 3 | 26 |
| C32 | 0.675 | 0.149 | 144 | 44 | 62 | 2791 | 650 | 384 | 506 | 5 | 51 |
| C33 | 0.684 | 0.399 | 100 | 20 | 32 | 392 | 306 | 206 | 645 | 3 | 20 |
| C34 | 0.695 | 0.332 | 851 | 13 | 31 | 2196 | 615 | 848 | 210 | 6 | 14 |
| C35 | 0.696 | 0.246 | 10 | 10 | 12 | 871 | 201 | 290 | 485 | 2 | 3 |
| C36 | 0.701 | 0.373 | 27 | 14 | 60 | 2098 | 833 | 244 | 2285 | 2 | 16 |
| C37 | 0.706 | 0.133 | 53 | 71 | 37 | 1322 | 489 | 302 | 1443 | 3 | 49 |
| C38 | 0.713 | 0.094 | 1796 | 9 | 18 | 5052 | 347 | 879 | 1168 | 4 | 14 |
| C39 | 0.72 | 0.304 | 1676 | 10 | 12 | 1118 | 1872 | 36 | 3155 | 3 | 11 |
| C40 | 0.726 | 0.348 | 389 | 41 | 33 | 1842 | 4402 | 598 | 2966 | 5 | 43 |
| C41 | 0.727 | 0.309 | 70 | 13 | 9 | 1223 | 548 | 241 | 3071 | 2 | 10 |
| C42 | 0.749 | 0.377 | 891 | 480 | 585 | 1891 | 1783 | 91 | 2138 | 6 | 344 |
| C43 | 0.765 | 0.19 | 2001 | 12 | 24 | 1519 | 259 | 473 | 3312 | 2 | 11 |
| C44 | 0.771 | 0.13 | 13 | 11 | 85 | 2403 | 1090 | 598 | 1536 | 3 | 16 |
| C45 | 0.781 | 0.131 | 1096 | 26 | 466 | 3722 | 1554 | 1781 | 1731 | 10 | 21 |
| C46 | 0.785 | 0.084 | 790 | 152 | 287 | 4108 | 5443 | 171 | 3325 | 4 | 258 |
| C47 | 0.791 | 0.205 | 330 | 11 | 18 | 3357 | 1197 | 183 | 1453 | 1 | 15 |
| C48 | 0.809 | 0.082 | 1546 | 648 | 33 | 1676 | 622 | 439 | 754 | 5 | 9 |
| C49 | 0.809 | 0.141 | 1478 | 80 | 166 | 3797 | 2331 | 960 | 1709 | 11 | 124 |
| C50 | 0.813 | 0.189 | 2329 | 180 | 459 | 307 | 1825 | 384 | 1959 | 3 | 335 |
| C51 | 0.815 | 0.372 | 140 | 39 | 104 | 1083 | 219 | 550 | 217 | 3 | 49 |
| C52 | 0.861 | 0.32 | 233 | 30 | 119 | 425 | 741 | 125 | 2896 | 9 | 30 |
| C53 | 0.883 | 0.122 | 19 | 45 | 41 | 214 | 204 | 187 | 623 | 2 | 48 |
| C54 | 0.898 | 0.161 | 701 | 10 | 19 | 2354 | 275 | 100 | 2070 | 5 | 20 |
| C55 | 0.961 | 0.321 | 979 | 75 | 144 | 2244 | 3249 | 1127 | 2419 | 2 | 65 |
| C56 | 0.976 | 0.194 | 2897 | 41 | 84 | 4761 | 2614 | 798 | 807 | 42 | 63 |
| C57 | 0.996 | 0.097 | 3252 | 837 | 353 | 816 | 731 | 244 | 1570 | 4 | 20 |

Samples consisted of 11 negative controls from the S protein ELISA, 11 that were high positive for both S and RBD protein in the ELISA, and 57 samples with a progressive increase in S protein OD value (2nd column) from the ELISA, but that were RBD negative. The Luminex threshold for positivity was set from the negative controls as the average plus 3 standard deviations for SARS-CoV-2 S protein = 134.05, RBD protein of = 73.63, and Nucleocapsid protein = 1088.50. Two columns showing data from the S protein and RBD protein ELISA are given for comparison to all the Luminex immunodetection data for S protein, RBD, Nucleocapsid, HCoV-229E S1, HCoV-HKU1 S1, HCoV-NL63 S1, HCoV-OC43 S1, MERS-CoV S1 and SARS-CoV-1 S1. The green boxes are values that were considered positive in the Luminex assay.

We also used the Luminex platform to assess correlation or cross-reactivity between donor antibody status for the S protein and positivity against the 4 human cold causing coronaviruses (HCoV-229E, -NL63, -OC43, and -HKU1). While many donors show antibodies against the 4 hCoVs in all three of our S protein ELISA groups (11 negatives, 11 high positives, and the 57 low positives), there was no correlation between antibodies against the 4-human cold-causing coronaviruses and antibody positivity or lack of positivity for SARS-CoV-2 S or RBD protein (Table 4). Thus, past infection status with the 4-human cold-causing coronaviruses did not impact S protein or RBD positivity.

Interestingly, of the 802 S protein ELISA positive donors, 108 donated blood or blood products at least 2 times from August to December 2020, and hence the study monitored maintenance or loss of antibody reactivity over time (Table 5). Analysis of these repeat donors showed 38 that had significant antibodies against S protein by ELISA on their first donation but that fell just below the assay threshold on the second donation, with an average time of 53 days (Table 5). In contrast, 24 donors maintained significant antibody reactivity and remained positive between the 2 donations, with an average time span of 69 days. However, of the 38 that lost positivity by the 2nd donation the initial composite ELISA OD value was 0.5960, while the group of 24 donors that maintained ELISA positivity between the 1st and 2nd donation had a much higher OD value of 1.47, which compares to a total average of 1.23 OD units across all 802 S protein positive donors (Table 5). Thus, the group of 38 repeat donors that lost their antibody positive status on the 2nd donation likely reflects the low starting point of antibody reactivity in conjunction with the known gradual loss of antibodies to SARS-CoV-2 over time (13), which now was below the sensitivity of the assay.

**Table 5.** Assessment of S raw values in donors that donated blood at least 2 times, some maintained positivity for spike protein antibodies, while some lost positivity.

|  | N | Mean Pos OD | Mean # of days | p value |
| --- | --- | --- | --- | --- |
| Pos-Neg | 38 | 0.596 | 52.8 |  |
| Stayed Pos | 24 | 1.466 | 69.3 | 0.0119* |
| All Positives | 802 | 1.228 |  |  |

p-value is for comparison of mean OD value of Pos-Neg donors compared with mean OD value of "Stayed-Pos" donors. The Pos-Neg donors were positive for S protein antibodies on first read only for the average number of days in between the 2 donations as shown. The "Stayed-Pos" donors had at least 2 positive S protein antibody readings over the time shown in days. The donors that lost positivity had a low first OD reading, while the donors that maintained positivity on their 2nd readings began with a higher initial S protein ELISA OD value.

Sample collection from August 13th - December 8th of 2020 was then split in half, which showed a significant increase in rates of positivity from the first to the second periods (Table 6). Specifically, rates of S protein ELISA positivity were 7.56% across the GCMA from August 13th through October 10th (58 days), compared with 9.24% from October 10th through December 8th, 2020 (58 days). Rates of S protein reactivity were also analyzed by month, which showed a temporal increase, culminating in a value of 12.86% in the portion of December that was evaluated (Table 6 and Figure 2). Thus, the GCMA emerged into the national vaccination phase of the pandemic with a background level of ≥13% of individuals likely with some degree of protection from SARS-CoV-2 re-infection.

**Table 6.** Rates of GCMA S protein antibody positivity broken into 2 successive time periods from August to December 2020.

|  | Number of Spike OD positive | N | % positive |  |
| --- | --- | --- | --- | --- |
| All | 802 | 9550 | 8.398 |  |
|  |  |  |  | p value |
| 1st 58 days | 362 | 4786 | 7.564 |  |
| 2nd 58 days | 440 | 4764 | 9.236 | 0.0016 |

The first 58 days spanned from August to October 2020, while the second 58-day period spanned from the rest of October to the first part of December 2020.

**Figure 2.**
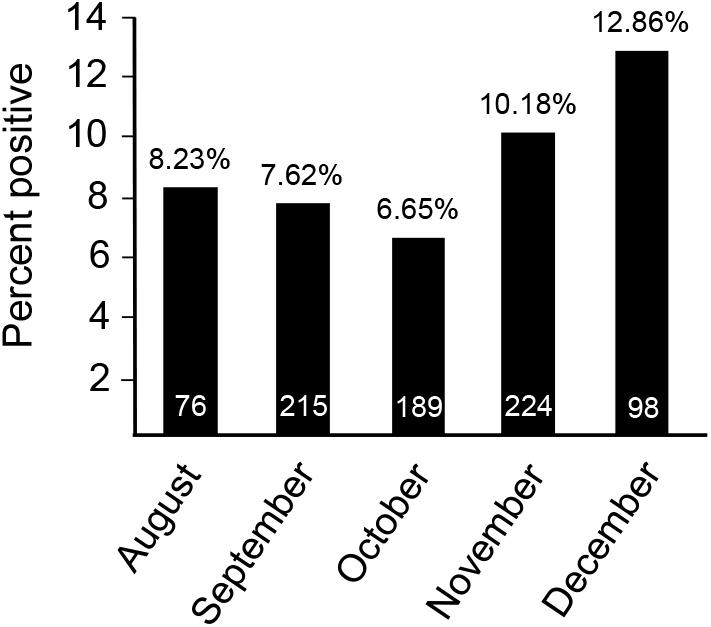
Percent positive S protein ELISA seroprevalence by month from August 13, 2020 to December 8, 2020. Month by month breakdown of the percent positive rate for 9550 unique donors. In August, 76 of 924 donors were positive (8.225%); September, 215 of 2821 donors were positive (7.621%); October, 189 of 2842 donors were positive (6.650%); November, 224 of 2201 donors were positive (10.177%); and December, 98 of 762 donors were positive (12.861%). p-value was determined from the z score for each month compared to the August positive rate for significance. Only December was significant as compared to August (z=-3.114, p=0.0018 Two-tailed) while September, October and November were not significantly different from each other. The number of samples collected per month is shown in each bar.

Within the donor population dataset, we also evaluated age ranges for rates of prior infection. The 9550 donors spanned in age from 16 to 91 years, and we analyzed rates of ELISA S protein positivity based roughly on decade increments. Interestingly, the youngest group of donors from 16-30 years of age had the highest rates of antibodies against S protein compared with individuals in their 30s and 40s, while adults over 60 years of age had significantly lower rates (Table 7). This significant reduction in rates in donors over 60 may reflect greater adherence to social protective guidelines or due to reduced antibody levels in older age groups, such that they more quickly lose responsiveness in the S protein-based ELISA. Finally, gender association with rates of S protein antibodies was 8.52% in males versus 8.28% in females, which was not statistically different (Table 7).

**Table 7.** Rates of S protein antibody positivity by ELISA from the indicated donor age ranges or by gender.

|  |  | Number of donors | Number of Spike OD positive | Percent positive | p value |
| --- | --- | --- | --- | --- | --- |
| Age group of donors | 16-30 | 1442 | 136 | 9.431 | 0.0286 |
|  | 31-40 | 1447 | 108 | 7.464 | n.s. |
|  | 41-50 | 1571 | 140 | 8.912 | n.s. |
|  | 51-60 | 2185 | 200 | 9.153 | n.s. |
|  | 60+ | 2905 | 218 | 6.984 | 0.0144 |
| Sex of donors | Male | 4564 | 389 | 8.523 |  |
|  | Female | 4986 | 413 | 8.283 | n.s. |

Only the 16-30 age range and the 60+ age range was statistically different from 31-40 age range, but the 31-40, 41-50 and 51-60 were not statistically different from each other. Rates in males and females were not significantly different.

Finally, we also analyzed geographic subregions within the GCMA for rates of S protein antibodies. Unfortunately, sampling was not large enough to examine rates based on individual zip codes, although statistical evaluation of the GCMA as larger subregions was possible, such as West versus East side, as split by Interstate 75 (I-75), and as rates in Ohio versus Kentucky, as split by the Ohio River (Table 8 and Figure 3). The data show a rate of 9.63% on the West side of Cincinnati versus 8.13% on the East side, while the Ohio portion of the GCMA was 8.79% versus 7.03% in the adjoining Kentucky region (Table 8 and Figure 3). Thus, the West side of Cincinnati had the highest rates of past SARS-CoV-2 infectivity, while the adjoining Kentucky region of the GCMA had the lowest.

**Table 8.** Rate of positive S antibody positivity clustered by regions within the GCMA that was tied to home address zip codes.

| Ohio* | Number of donors | Number of positive donors | Percent positive | p value |
| --- | --- | --- | --- | --- |
| West of I-75 | 3136 | 302 | 9.630 |  |
| East of I-75 | 4266 | 347 | 8.134 | 0.0123 |
| Ohio | 7418 | 652 | 8.789 |  |
| Kentucky | 2132 | 150 | 7.036 | 0.005 |

\*Excludes Kentucky and 17 people (3 positive) in zip codes divided evenly by I-75. However, Ohio was also summated along with the additional 17 donors on the I-75 border to compare against Kentucky zip codes on the south side of the Ohio River, but within the GCMA.

**Figure 3.**
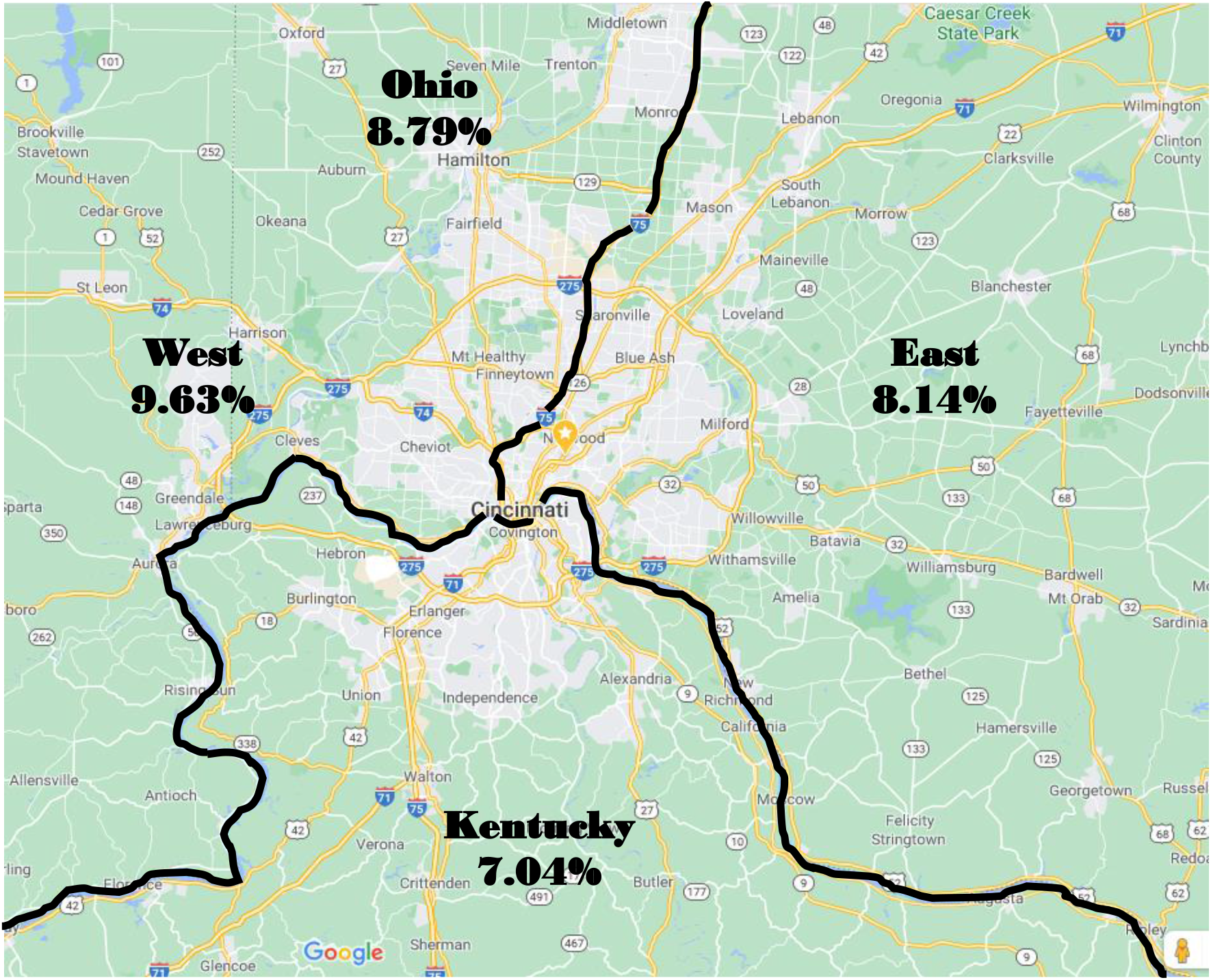
Seroprevalence of protein antibodies in the Greater Cincinnati Metropolitan Area. Map image courtesy of Google Maps (https://www.google.com/maps). Greater Cincinnati Metropolitan Area (GCMA) as defined by the United States Census Bureau (https://data.census.gov/cedsci/map?q=All%20counties%20in%20Ohio&g=310M500US17140&tid=ACSDP5Y2019.DP02&layer=VT_2019_310_M5_PY_D1&cid=DP02_0093PE&palette=Teal&break=5&classification=Natural%20Breaks&mode=customize) and reported in the 2010 AGE, RACIAL, GENDER AND MARITAL STRUCTURE OF GREATER CINCINNATI. The Cincinnati Metropolitan Statistical Area (MSA) includes: Butler, Brown, Clermont, Hamilton, and Warren Counties in Ohio; Boone, Bracken, Campbell, Gallatin, Grant, Kenton, and Pendleton Counties in Kentucky; and Dearborn, Franklin, and Ohio Counties in Indiana. The indicated data show percent of seropositivity for Spike antibodies shown by the indicated regions.

## Discussion

To our knowledge the current study is the first to report rates of SARS-CoV-2 seroprevalence in the GCMA immediately preceding the national vaccination program. The total cumulative rate as of December 2020 was ∼13%, which should help achieve herd immunity in the GMCA more rapidly in conjunction with the national vaccination program (14,15). With respect to immunity after viral infection, 150,000 previously infected individuals in Ohio and Florida were tracked from March 2020 -August 2020 and shown to be largely resistant to subsequent re-infection, like protection achieved with vaccination (6). In the current study here, the highest rates of past infection were observed in individuals under 30 years of age, and more generally on the West side of the GCMA compared to the East side and adjoining regions of Kentucky. Data trends failed to reveal a difference in background levels of past infectivity based on ethnicity in the GCMA as analyzed in Caucasians, African Americans, Hispanics, and Asians, although the total sampling pool of the later 3 ethnic groups was too low to achieve statistical certainty.

The detection by seroprevalence of past infection of SARS-CoV-2 is dependent on the quantitative measures of the S protein-based IgG-dependent ELISA. Thus, exact details surrounding implementation and associated quality control measures are critical in achieving accuracy of past purported infectivity rates within the population. A modified ELISA protocol from Mount Sinai Icahn School of Medicine in New York City (9,10) was implemented, which was given an Emergency Use Authorization (EUA) by the US Food and Drug Administration (FDA) in April of 2020 (16). Additional quality control measures included the use of 60 serum samples obtained prior to the onset of the pandemic as true negatives in generating a background value for the S protein ELISA. The entire ELISA process was also automated with robotic liquid handling and plate reading systems. Hence, the values reported here are accurate in representing the rates of past infectivity in the donor samples evaluated within the GCMA. However, it is likely that the ELISA testing platform used here will miss individuals in whom the levels of antibody have dropped below the level of detection, as previously reported (17). Thus, not all individuals infected in the early months of the pandemic will maintain a positive reading in this ELISA and as such, the composite rate calculated from the S protein ELISA likely underestimates the true rates of past infectivity in the GCMA, which is likely several percentage points higher (see below).

Individuals who present to donate blood or other blood products are not a true cross-sectional representation of a metropolitan area. Indeed, such individuals are pre-screened for communicable diseases or behaviors that are high risk for attaining such diseases. The ethnicity of the GCMA blood donor volunteers, especially during the current pandemic, is 90% or greater Caucasian and hence, under-represented in other ethnicities such as African Americans and Hispanics (18). Moreover, blood donors also tend to be healthy and have lower prevalence of acquired chronic health conditions. However, given that this study was a post-hoc analysis of de-identified specimens, no selection bias based on donor suspicions of past infection were involved. Indeed, as discussed below, the rates of seropositivity observed here are in line with rates calculated by PCR analysis across the entire state of Ohio since the inception of the pandemic (3).

In the past year countless seroprevalence studies have been published or uploaded to preprint servers from across the USA, although very few have thus far extended to December 2020. A few of these past studies are particularly relevant and interesting to consider in relation to the current analysis. One such study examined 252,882 blood donors over 24 centers across the USA from the months of June and July 2020 (19). Vassallo et al., utilized the Ortho VITROS Anti-SARS-CoV-2 total immunoglobulin assay for the S1 region of the S protein for IgG, IgA and IgM (19), which was different from the full-length S protein ELISA detecting IgG serum levels that was produced in house and employed here. Vassallo et al., reported a rate of 1.83% in June and 2.26% in July across their entire USA sample population. Within these data were results from Chicago, which showed a rate of 2.76% in June and 3.34% in July (19). By comparison, another seroprevalence study from the Chicago area that analyzed 1545 solicited volunteers, showed a seroprevalence of 19.8% from June 24 through September 6, 2020 (20). This later study utilized the same S protein ELISA protocol from Krammer and colleagues (9,10) that we also used to investigate the GCMA, although their volunteer sample collection was based on advertising for serology evaluation (20). Hence, select community-based recruitment variables and the technical aspects of the immunodetection platform could underlie the widely disparate results discussed here. However, based on PCR measured SARS-CoV-2 molecular detection from the beginning of the pandemic until March of 2021, there were approximately 980,000 Ohioans infected with the virus. This equates to a rate of 8.23% of the state’s population, which is generally consistent the data showing a rate of ∼12% seropositivity as of December of 2020 in the GCMA. More interestingly, extrapolation of these data 3 additional months to the time this report was uploaded in March 2021, the rate of past infectivity in the GCMA is likely ∼16%, and as stated above this approximation almost certainly slightly under-estimates the true rate due to the known gradual decline in antibody levels and the associated drop below the threshold of the S protein-based ELISA (13,17). Given all these factors and the fact that blood donors likely represent a slightly healthier cross-section of the GCMA, an overall estimated rate of past infection of roughly 20% seems reasonable for the present day in mid-March 2021.

Another study examined 177,919 seemingly random adult blood samples from across 50 States that spanned from July 27 through September 24, which reported a range of values from just under 1% to over 20%. This study used 3 different automated clinical laboratory immunodetection platforms for either S or N protein. Within their data set samples collected from August 24 - September 24, 2020 overlap with part of our collection time, and in Ohio they report values of 2.8 to 5.0% (21). This value is also consistent with another more limited seroprevalence study with 727 samples from across the entire state of Ohio in July 2020, which incorporated mathematical modeling to predict a rate of 7% past viral infectivity (22), a value that is close to our actual data from the adjacent month of August in the GCMA.

While there are a multitude of seroprevalence studies in the literature from across the USA, only a handful appeared directly relevant to the interpretation of this study. These past studies generally support our conclusions and suggest that the ELISA implemented here was rigorous and properly calibrated, and that the donor subgroup used generated a reasonable approximation of the GCMA and likely the entire state of Ohio. The data in this study establish a rate of ∼13% past SARS-CoV-2 infectivity within the GMCA by the end of 2020, and extrapolation to the present day (March of 2021) approximates a rate of 16% past infectivity, and likely even as high as 20% if depreciation in blood antibody levels over time is considered. This knowledge can impact the deployment of the vaccination program to most rapidly achieve herd immunity (15). For example, previously infected individuals might only need 1 vaccine dosage for full protection compared with a naïve individual who requires 2 vaccinations (with Moderna and Pfizer vaccines). Indeed, previously infected and recovered individuals produce a strong immunologic reaction after a single dosage of the Pfizer vaccine that is comparable to the standard 2 dose routine in naïve individuals (23); and using this information and associated strategy would augment the relative supply of the vaccine in attempting to achieve herd immunity more rapidly.

## Methods

### Human Samples

Blood samples were collected in EDTA tubes from volunteer donors presenting to the Hoxworth Blood Center according to USA FDA regulations and American Association of Blood Banks (AABB) guidelines. Specimens were de-identified and as such, the University of Cincinnati Institutional Review Board (FWA #: 000003152) ruled that these blood donor samples and their analysis as constituting non-human research for the proposed study of SARS-CoV-2 serological responsiveness. Donors are subjected to medical, social, behavioral, and travel history questionnaire to reduce risk of communicable diseases in the donated blood following USA FDA regulations and AABB guidelines. Donors who felt unhealthy including, but not limited to, elevated temperature, low blood hematocrit or signs of respiratory infection were excluded. However, donors were not questioned as to their history of SARS-CoV-2 infection, if they were symptom-free, and were not tested at the time of collection. Even though donors were de-identified, repeat donations from the same donor were obvious based on date of birth, zip code, blood group type, and gender data.

### ELISA

The ELISA protocol was adopted from 2 extensively annotated reports in the literature (9,10). Briefly, SARS-CoV-2 antigens for S protein and RBD were coated on 96 well plates (Corning 9018) in 1X PBS (Fisher) and stored between 1 to 7 days at 4 °C. S protein was coated at 1.0 µg/ml in 50 µl per well and the RBD protein fragment was coated at 2 µg/ml in 50 µl per well. The next day antigen plates were washed 5 times with 1X PBS + 0.1% Tween-20 (PBST) and then blotted so that the wells have no residual volume but are not technically dry. Blocking was performed with 3% non-fat dry milk (NFDM) in PBST for 1 hour at room temperature. Antigen plates were again washed 5 times with PBST. Samples received from Hoxworth Blood Center in EDTA anticoagulated tubes were heat inactivated at 56°C for 20 minutes, centrifuged and the resulting plasma was diluted 1:10 in 1X PBS and stored at −10°C until use. On day of use, samples were again diluted 1:10 in PBST + 1% NFDM. Controls on each plate consisted of a plasma sample with known high S protein antibody levels. Exactly 50 µl of the 1:100 diluted samples were added to 96-well blocked antigen plates described above (9,10). The plasma samples were allowed to incubate in the 96-well antigen plates for 2 hours at room temperature and then washed 5 times using a BioTek plate washer ELx405. Plates were blotted to remove all liquid and then 50 µl of goat anti-human IgG conjugated to horseradish peroxidase (HRP) (Jackson Labs 109-035-008) in PBST was added at a dilution of 1:10,000 for 1 hour at room temperature. Plates were washed 5 times with PBST and once with citric acid phosphate buffer, pH 5.0. The colorimetric reagent specific for HRP activity assessment, OPD (Sigma P4664), was added in water to the plates for 15 minutes at room temperature and the reaction was stopped with the addition of 1 M H_2_SO_4_. Spectrophotometric based absorbance at 492 nm was assayed in the BioTek Synergy 2. Negative control serum samples from 60 individuals were used to establish the absolute baseline value for the S protein ELISA and 53 individuals for the RBD protein ELISA, and 3 times the standard deviation was summated to this average negative value in assigning a positive value threshold.

### Protein Production and Purification

Two different versions of the S protein were generated based on expression constructs received from Krammer and colleagues (9,10). The first construct expresses a full length trimeric and stabilized version of the S protein and the second was the much smaller RBD from within the primary sequence of the S protein. The sequence used for both proteins is based on the genomic sequence of the first virus isolate, Wuhan-Hu-1, released on January 10th, 2020, which was optimized for codon usage and mutated to remove the polybasic cleavage site, as well as addition of 2 mutations to stabilize the protein (9,10). At amino acid P1213 the sequence was also fused to a thrombin cleavage site, a T4 foldon sequence for proper trimerization and a C-terminal hexahistidine tag for purification. The sequence was cloned into a pCAGGS vector for expression in mammalian cells (9,10).

RBD and S proteins were produced by transient transfection of expiCHO™ cells (ThermoFisher, A29133) via manufacturer’s instructions. Briefly, expiCHO cells were transfected with plasmid DNA (1 µg/ml of cell volume) at 6×10^6^ cells/ml in suspension culture using the Expifectamine reagent. Transfected expiCHO cells are then cultured per the manufacturers ‘max titer’ protocol at 32 degrees shaking at 125 rpm for twelve days. Cell culture supernatants were harvested and filtered through a 0.2 μM membrane and both S protein and RBD were purified using a 20 mL Ni^2+^-charged HiPrep IMAC FF 16/10 column (Cytiva) to bind the His-tagged region engineered into each protein (9,10). The binding buffer was composed of 20 mM Tris pH 7.4, 500 mM NaCl, and 10 mM imidazole. After loading the protein onto the column, a wash was performed using 3 column volumes of binding buffer. The protein was eluted with a step gradient to 20 mM Tris pH 7.4, 500 mM NaCl, 300 mM imidazole. Fractions from the elution were buffer-exchanged into 1X PBS using a HiPrep 26/10 Desalting column (Cytiva, GE17-5087-01). For RBD, the buffer-exchanged protein was further purified using a HiLoad 26/600 Superdex 75 pg (Cytiva, GE28989334) 320 mL column with 1X PBS as the running buffer. A 10 kDa MWCO centrifugal filter unit (Amicon, ACS501024) was used to concentrate fractions containing RBD. Protein purity was validated by SDS-PAGE and western blotting using a PENTA-his antibody (Qiagen, ID:34660). Quantification of protein was performed using a BioMate 3S spectrophotometer (Thermo Scientific). The ProtParam web server was used to calculate the molecular weight (RBD: 27,496.24 Da; Spike: 139,195.53 Da) and extinction coefficient at 280 nm (RBD: 33850 M^-1^ cm^-1^; Spike: 142835 M^-1^ cm^-1^) based on the amino acid sequences of RBD and S. Purified protein was stored at −20°C in 50% glycerol with 5 mM sodium azide.

### Analytical ultracentrifugation

Purified RBD and S proteins were characterized by sedimentation velocity analytical ultracentrifugation using a Beckman Coulter XL-I. RBD samples after Ni-NTA or gel filtration chromatography were dialyzed as needed into 1X PBS and spun at 48,000 rpm at 20°C in an An-60 Ti rotor. Purified spike protein from a −20°C aliquot was buffer exchanged into 1X PBS using a HiPrep 26/10 desalting column as mentioned above. The samples were then spun at 32,000 rpm at 20°C using meniscus-matching centerpieces (Spin Analytical, Inc., South Berwick, ME). Data were analyzed using SEDFIT’s continuous c(s) distribution model (24), SEDANAL version 7.45 (25), or DCDT+ version 2.4.3 (26).

### Luminex

Luminex assays were performed with the One Lambda COVID Plus kit according to manufacturer’s instructions (Themo Fisher, LSCOV01). Briefly, the diluted plasma/serum samples and controls from the ELISA screen were combined in a 96-well MultiScreen filter plate (EDM Millipore, MSVN1B50), 2 µl of serum/plasma was added to 17 µl of 1X PBS and then 1 µl of 0.02 M EDTA was added for a total volume of 20 µl (final serum dilution 1:10). According to the kit the beads were prepared by vortexing and then 5 µl of this bead mix was added to each sample well. The plates were sealed and incubated for 30 min in the dark on a plate shaker at room temperature. After incubation, the seal was removed and washed with 150 µl of kit wash buffer in each well. The contents of each well were then removed with a standard vacuum manifold (EMD Millipore, #40-097) as part of the washing procedure. The plates were washed 2 more times with 150 µl of wash buffer, then 100 µl of PE-conjugated anti-human IgG (One Lambda, LS-AB2) was added at a 1:100 and incubated for 30 min. The plates were then washed 3 times with 150 µl of wash buffer per well and then 80 µl of 1X PBS was added and incubated for 5 minutes on a plate shaker. The plates were loaded into the Milliplex 200 with reporter laser 532nm / and classification laser 635nm for analysis (Luminex). Data from the instrument were prepared and analyzed using Bio-plex manager 6.1 software.

### Statistics

Means and Standard Deviations were determined for all data sets using Microsoft Excel Data Analysis Descriptive Statistics tool; histograms were made using the Microsoft Excel Data Analysis Histogram tool and all graphs were made using Microsoft Excel. Statistics between groups were calculated using the Microsoft Excel Data Analysis t-Test: Two-Sample Assuming Unequal Variances. All z scores and the number of standard deviations from the mean of the reference population were calculated for the difference between rates of two data sets and subsequent p-value calculated using Microsoft Excel. Microsoft Excel Data Analysis Correlation tool was used to produce correlations within the Luminex data shown in Table 4.

## Supporting information

Conflict of interest Disclosure

Reporting checklist

## Data Availability

Requests datasets or protocols used in this study should be directed to the corresponding author and will be made available to investigators upon reasonable request.

## Acknowledgements

We would like to thank Dr Florian Krammer at Mount Sinai Icahn School of Medicine for providing the mammalian expression vectors for the modified S protein and the RBD protein (9,10), upon which the ELISA platform was generated in house.

## Ethics and Reporting

Blood samples were collected from volunteer donors presenting to the Hoxworth Blood Center following United States FDA regulations and American Association of Blood Banks (AABB) guidelines with a signed standard donor consent form. Specimens were de-identified and as such, the University of Cincinnati Institutional Review Board (FWA #: 000003152) ruled that these blood donor samples and their analysis as constituting non-human research for the proposed study of SARS-CoV-2 serological responsiveness.

