## Supplemental Figure 1 for "Seroprevalence of SARS-CoV-2 Infection in Cincinnati Ohio USA from August to December 2020"

**Supplemental Figure 1.** Expression, purification, and characterization of SARS-CoV-2 S and RBD proteins. A) Comparison of expression yields in different cell lines. Dashed lines represent the reported yields from Stadlbauer et al. (10). B) Reducing SDS-PAGE gel showing RBD purified by Ni-NTA and gel filtration chromatography and Spike protein purified by Ni-NTA chromatography. C-E) Sedimentation velocity analytical ultracentrifugation analysis of protein quality and assembly state in solution. C) Sedimentation coefficient distribution for RBD purified by Ni-NTA revealing monomer and disulfide-linked dimer species. D) Sedimentation coefficient distribution for monomeric RBD (experimental MW of 31.1 kDa) purified by S75 gel filtration. E) Sedimentation coefficient distribution for Spike protein showing that trimer is the predominant species (experimental MW of ~519 kDa). The trimer sedimentation coefficient of 12.9 S was consistent with the value (12.6 S) calculated by HullRad (27) hydrodynamic modeling of the glycosylated spike trimer structure (PDB 6VXX).

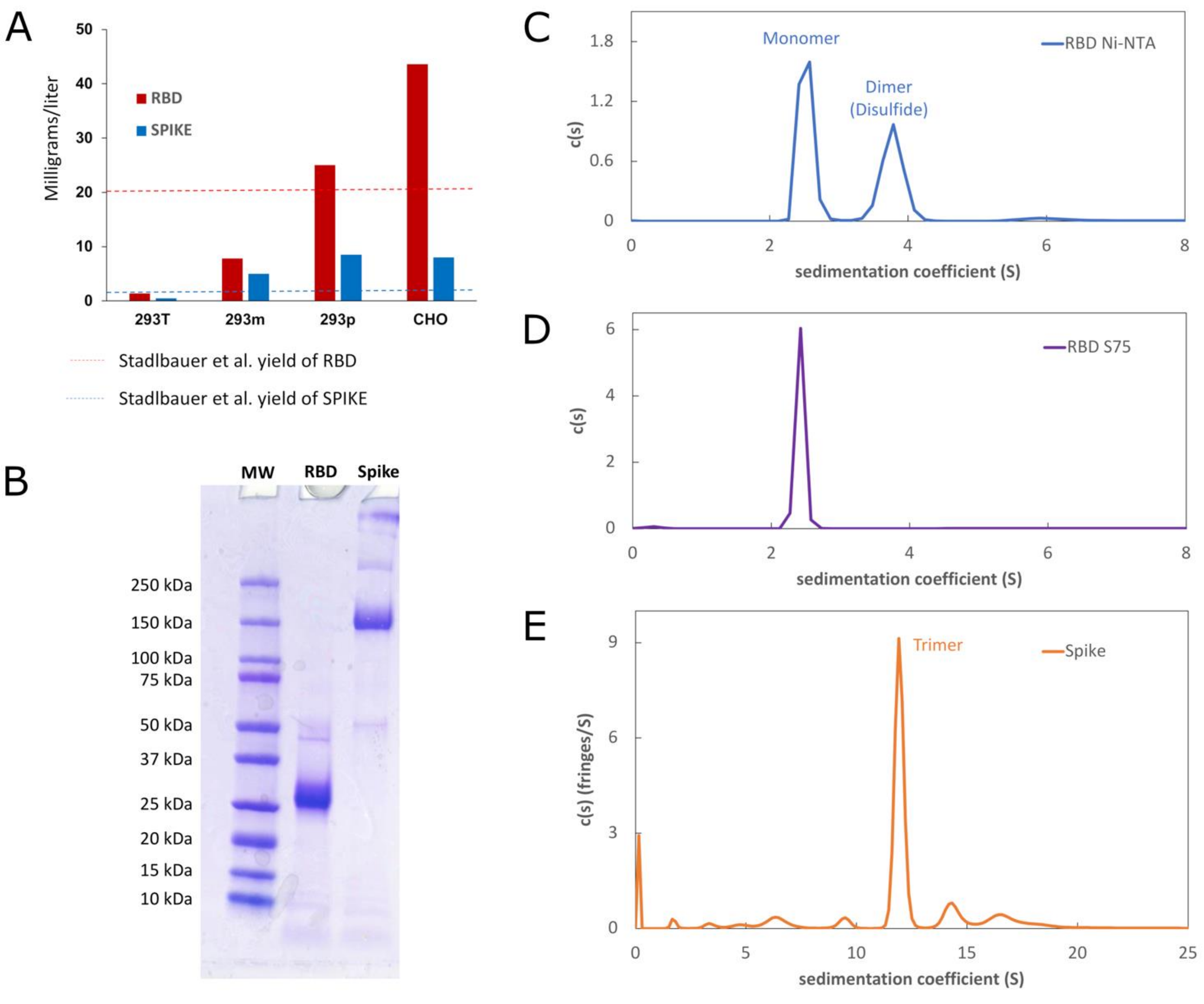
